# Could Deficiencies in South African Data Be the Explanation for Its Early SARS-CoV-2 Peak?

**DOI:** 10.1101/2020.08.31.20185108

**Authors:** S. J. Childs

## Abstract

The SARS-CoV-2 pandemic peaked very early in comparison to the thresholds predicted by an analysis of prior lockdown regimes. The most convenient explanation is that some, external factor changed the value of the basic reproduction number, r_0_; and there certainly are arguments for this. Other factors could, nonetheless, have played a role. This research attempts to reconcile the observed peak with the thresholds predicted by lockdown regimes similar to the one in force at the time. It contemplates the effect of two, different, hypothetical errors in the data: The first is that the true level of infection has been underestimated by a multiplicative factor, while the second is that of an imperceptible, pre-existing, immune fraction of the population. While it is shown that it certainly is possible to manufacture the perception of an early peak as extreme as the one observed, solely by way of these two phenomena, the values need to be fairly high. The phenomena would not, by any measure, be insignificant. It also remains an inescapable fact that the early peak in infections coincided with a fairly profound change in r_0_; in all the contemplated scenarios of data-deficiency.

## 1 Introduction

On around the 18th of July, 2020, the incidence of SARS-CoV-2 cases peaked and the number of active infections followed suit around five days later. Either some unknown factor caused the basic reproduction number, *r*_0_, to drop below unity, around the 13th of July (the mean SARS-CoV-2 incubation period is 5.2 days [14]), or the threshold had been reached. All this transpired at an apparently very low level of total infection, a mere 0.7 % of the population; or, couching this in the more conventional terms of susceptibility, 99.3 %. Such a threshold would imply a basic reproductive number less than 1.01. On the 13th of July, South Africa did revert to a lockdown regime similar to its previous level 4 lockdown (*r*_0_ = 1.69), referred to in this work as “level 3.5”, however, as much as prohibition, curfews and a number of other measures were reinstated, the threshold predicted by an analysis of the level 4 lockdown regime, suggests that a new regime, in itself, was not enough to cause the observed peak. A basic reproductive number of 1.01 is exceptionally marginal. How does one explain this conundrum?

It is already known that the perception of a peak at 99.3 % is based on infection-data which are deficient by an order of magnitude, or even greater. The head of the CDC, Robert Redfield’s opinion on the topic of asymptomatic or undiagnosed SARS-CoV-2 infections, in the U.S.A., is that antibody testing reveals that “A good rough estimate now is 10 to 1” ([5]) and others, in similar positions all over the world, have expressed similar sentiments. Redfield’s factor of eleven also needs to be revised upward if one considers that, although antibodies lend themselves favourably to the diagnosis of immunity, they are not the ultimate indicator. Undetected, T-cell mediated immunity can exist in the absence of a positive antibody test. In South Africa, epidemiologists have focussed on excess deaths and put forward a value of 1.59 ([10], [4] and [3]). There is not necessarily any conflict between this apparently, relatively low number of excess deaths and Redfield’s statement, if one considers the obvious bias in detection: If you’re so sick that you’re about to die, you’re more likely to seek out medical assistance and be diagnosed. It may also be worth keeping in mind that a massive 57 % of the inhabitants of the Mumbai slum areas of Chembur, Matunga and Dahisar tested positive for exposure to SARS-CoV-2 [1]. That the SARS-CoV-2 virus is often insidious and infection data are consequently incorrect by a factor is therefore already a widely recognised phenomenon. This fact, alone, is nonetheless unable to reasonably explain the SARS-CoV-2 threshold observed in the South African data, without contemplating improbably-high, though not impossible, values.

The question then arises as to whether the SARS-CoV-2 virus really is novel and the population really is naive, or whether some other pathogen, genetics, etc. has not imparted an undetectable immunity to a significant fraction of the population. Although antibodies lend themselves favourably to the diagnosis of immunity, they are not the ultimate indicator. As already stated, undetected, T-cell mediated immunity can exist in the absence of a positive antibody test.

This research attempts to reconcile the observed peak in South African infections with the thresholds predicted by lockdown regimes similar to the one in force at the time. It contemplates the effect of two, different, hypothetical errors in the data: The first is that the true level of infection has been underestimated by a multiplicative factor, denoted *a*, while the second is that of an imperceptible, pre-existing, immune fraction of the population, denoted *b*. Since the rules in place at the time of the peak were most similar to level 4, the values of *a* and *b* explored were mostly selected on the basis that they manufacture an erroneously-detected, 99.3 % threshold for the level 4 lockdown.

Of course, it is possible that some, other, external factor, caused the basic reproduction number to plummet, around the 13th of July, and a quantification of lockdown regimes means nothing if the public at large were not compliant with the rules.

## 2 The Erroneous Perception Created by Inexplicable Immunity and Asymptomatic, or Undiagnosed, Infections

Suppose that *S*(*t*), *I*(*t*) and *R*(*t*) represent the usual quantities in Kermack and McKendrick’s SIR model [13]. Suppose that a tilde is used to further distinguish detected values of these quantities, which for the purposes of this exposition, will be erroneous. If, for some presently inexplicable reason, there is an imperceptible, pre-existing immune fraction of the population, *b*, then 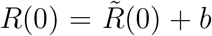, for an epidemic that begins at some time, *t* = 0. Suppose one were to further determine that both *Ĩ*(*t*) and therefore, the resistant portion arising from the current epidemic, 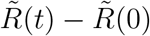, are in actual fact higher by some factor, *a*. That is,

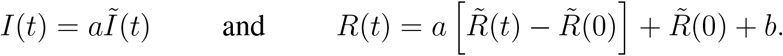

Substituting these quantities into

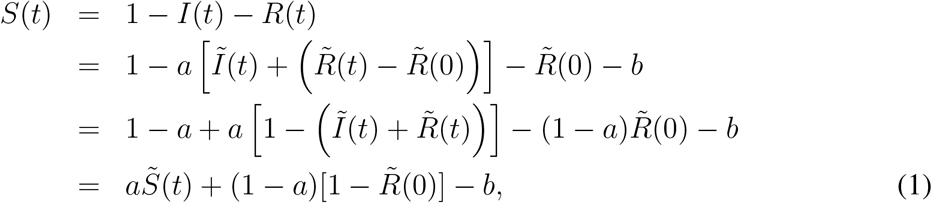

in which 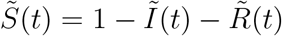.

In the particular case of the South African, SARS-CoV-2 peak, 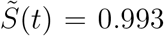 and 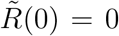. Equation (1) therefore simplifies to

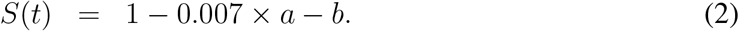

The true value of the erroneously-detected 99.3 % threshold therefore depends on the values of both *a* and *b*, some examples of which are given in Table 1.

**Table 1:** Some examples of what the true value of an erroneously-detected peak of 99.3 % would be, based on the substitution of various values of *a* and *b* into Equation (2).

| Detected Threshold, $\tilde{S} / \%$ | $a$ | $b$ | True Value, $S / \%$ |
| --- | --- | --- | --- |
| 99.3 | 1.0 | 0.1 | 89.3 |
| " | 10 | 0.0 | 93.0 |
| " | 10 | 0.83 | 10.0 |
| " | 20 | 0.65 | 21.0 |
| " | 30 | 0.5 | 21.0 |
| " | 60 | 0.0 | 58.0 |

Changing the subject of Equation (1),

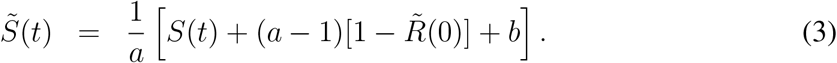

This is a formula for the perceived susceptible fraction that will be erroneously detected for given values of *a* and *b*.

## 3 The Data and Their Interpretation

Epidemiological data are usually presented in the format “numbers of current infections” and “total number of cases”. The present case of the SARS-CoV-2 pandemic is no exception. The data for the level-5 lockdown, the level-4 lockdown, the early level-3 lockdown and Sweden were already interpreted in [12] and may be found in Table 2. The final level-3 and so-called level-3.5 values, in Table 2, were determined as follows.

**Table 2:** The scenario in which the data is assumed to be correct. The basic reproduction number, *r*_0_, the consequent threshold and the point at which all infection ceases, *S*_*∞*_, have all been calculated with *a* = 1, *b* = 0. With the exception of Sweden, all are calculated from data associated with the relevant lockdown regimes, imposed for the SARS-CoV-2 pandemic, in South Africa.

| Regime | Date 1 | Total Cases | Active Cases | Date 2 | Total Cases | Active Cases | $r_0$ | Threshold $S$ / % | $S_\infty$ / % |
| --- | --- | --- | --- | --- | --- | --- | --- | --- | --- |
| level 5 | 10.04 | 1 940 | 1 589 | 01.05 | 5 945 | 3 518 | 1.93 | 51.8 | 22.3 |
| level 4 | 16.05 | 14 695 | 8 096 | 01.06 | 34 376 | 16 110 | 1.69 | 59.3 | 31.4 |
| level 3 | 16.06 | 76 210 | 32 580 | 01.07 | 159 617 | 803 30 | 2.34 | 42.7 | 13.0 |
| " | 16.06 | 75 493 | 30 996 | 13.07 | 289 922 | 146 472 | 2.17 | 46.0 | 16.2 |
| level 3.5 | 28.07 | 462283 | 164 680 | 18.08 | 591 132 | 94 098 | 0.65 | — | — |
| Sweden | 26.03 | 3 580 | 3 443 | 05.06 | 43 443 | 30 663 | 3.16 | 31.6 | 5.0 |

### The Level-3 Lockdown

The level-3 lockdown commenced on the 1st of June and was still in force up until the 12th of July. Once again, a period of 15 days was allowed for the viral incubation period and the subsequent diagnosis of an infection. Once again, it was also assumed that the termination of the level-3 lockdown would not reflect in the data for at least 24 hours. Curves were accordingly fitted to the subset of data ([7], [6] and [2]) which commenced on the 16 of June and terminated on the 13th of July. The curves fitted to the data, using Gnuplot, are depicted in Figure 1.

**Figure 1:**
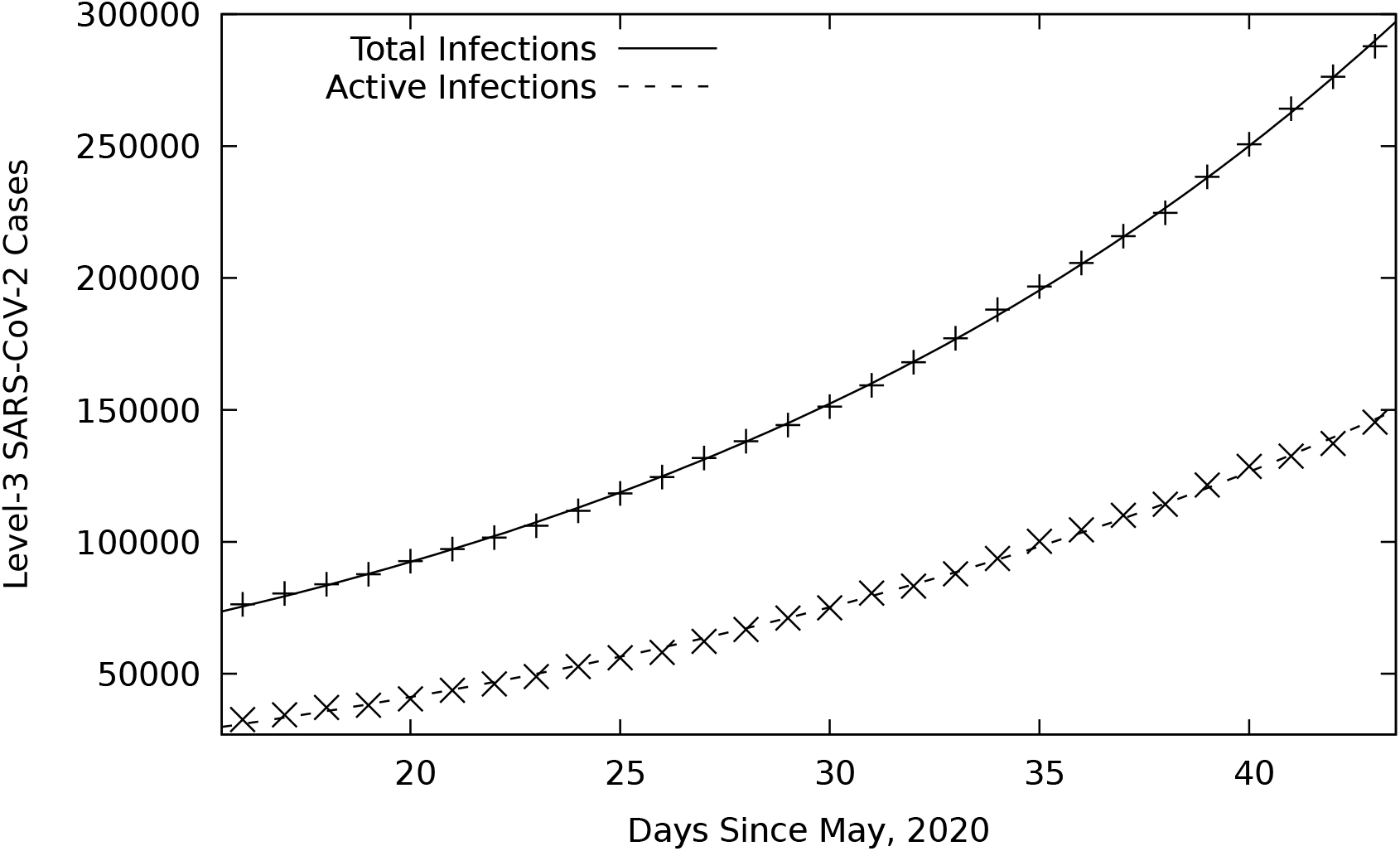
Level-3, best fits to SARS-CoV-2, infection data (16th of June to the 13th of July, 2020).

The formula for the “total infections” curve is

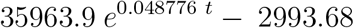

and the formula for the “active infections” curve is

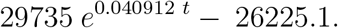

The values these formulae yield for the relevant dates are provided in Table 2.

### The Level-3.5 Lockdown

The so-called level-3.5 lockdown commenced on the 13th of July and was in force up until the 17th of August. Once again, a period of 15 days was allowed for the viral incubation period and the subsequent diagnosis of an infection. Once again, it was also assumed that the termination of the level-3 lockdown would not reflect in the data for at least 24 hours. Curves were accordingly fitted to the subset of data ([7], [6] and [2]) which commenced on the 28th of July and terminated on the 18th of August. The curves fitted to the data, using Gnuplot, are depicted in Figure 2.

**Figure 2:**
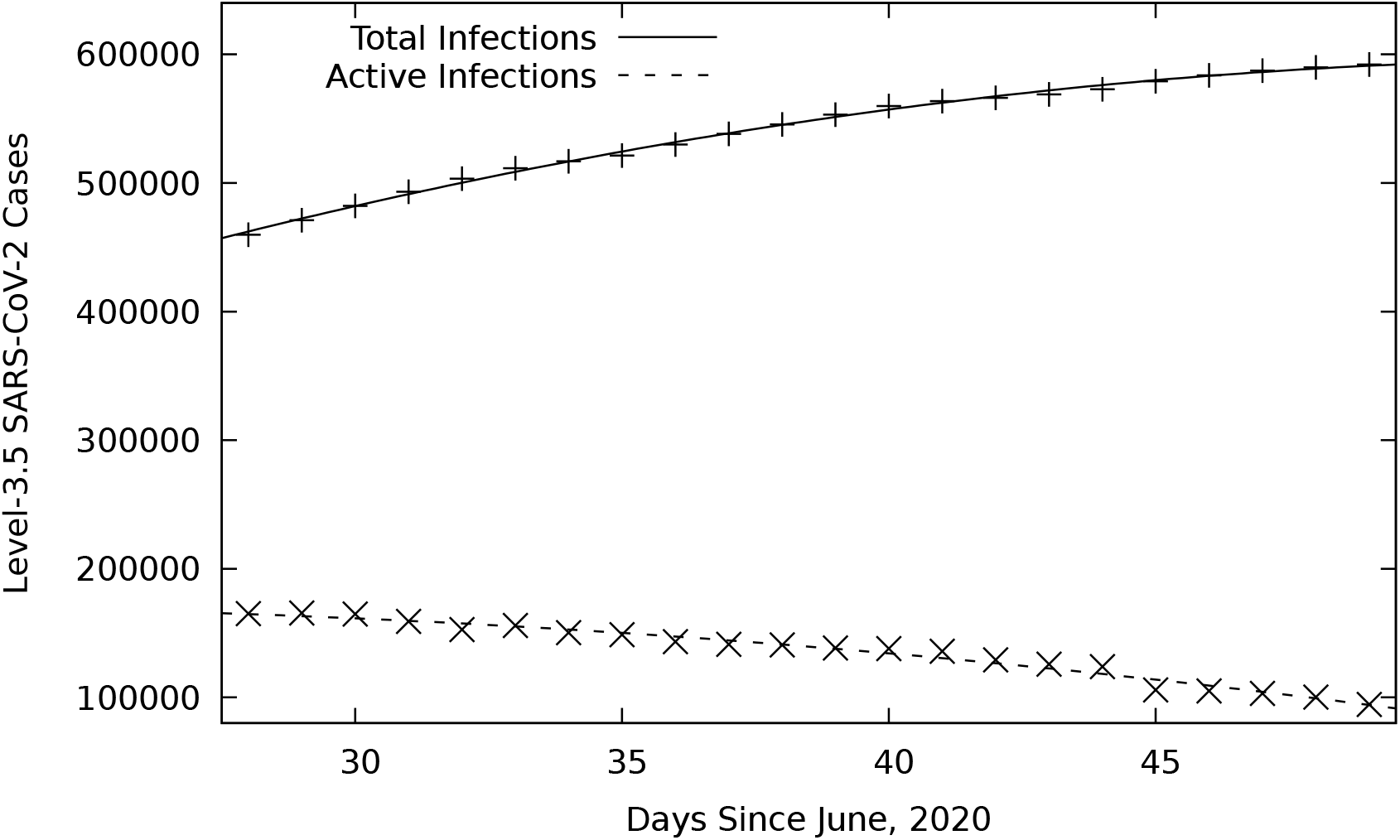
Level-3.5, best fits to SARS-CoV-2, infection data (28th of July to the 18th of August, 2020).

The formula for the “total infections” curve is

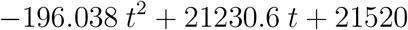

and the formula for the “active infections” curve is

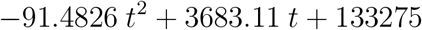

The values these formulae yield for the relevant dates are provided in Table 2.

### Converting the Conventional, Epidemiological Format into *S*(*t*) and *I*(*t*)

The epidemiological data have been presented in their usual format “numbers of current infections” and “total number of cases”. If *N* is the size of the population, “active infections” are just *Ĩ*(*t*)*N* and “total infections” are just 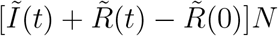. Realising this, the values of *S*(*t*) and *I*(*t*) needed for this work can be obtained from

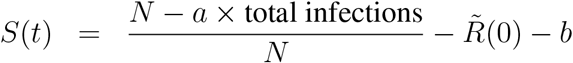

and

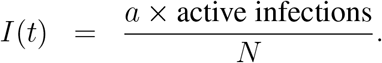

In 2020, the size of the South African population was estimated to be 59 140 502 by [8].

## 4 Calculation of the Basic Reproduction Number, *r*_0_

The basic reproduction number, *r*_0_, was calculated according to the formula derived in [12] from Kermack and McKendrick’s SIR equations [13]. That is,

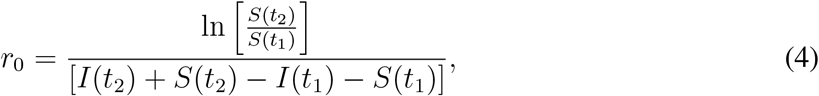

in which *S*(*t*), *I*(*t*) and *R*(*t*) represent the usual quantities in Kermack and McKendrick’s SIR model [13] and they are evaluated at either end of an interval, [*t*_1_, *t*_2_], in the formula.

Expedience was the motivation for using this slightly unorthodox method, however, not only have so-called individual level models, such as those of [9] and [11], been discredited by [11] as a means for calculating *r*_0_, they are also far more laborious than the simple method used in this work.

Notice that this formula also appears to be fairly robust. Take any movement of *I*(*t*) by an additive constant, up or down, for example. Such movement has no effect on the calculation of *r*_0_, whatsoever. This is important as the infection data, more than any other, are often plagued by exactly this problem. In fact, the formula, Equation (4), is reasonably robust against any data error that does not effect the relative values, or slopes of the functions concerned. This will be borne out when exploring the use of a multiplicative factor on the data.

## 5 Calculation of the Threshold and *S*_*∞*_

It is instructive to know both the threshold, as well as the point at which all infection would cease. Both are calculated from *r*_0_ and the relevant theory is provided in [12]. If the susceptible fraction of the population is still above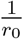, for a given regime, the epidemic will continue to grow in the way of infections. Only at the threshold does *r*-effective drop to unity and the infections subsequently decline. Once *r*_0_ has been calculated, *S*(*t*_2_) = *S*_*∞*_, can be recovered from Equation (4), by considering that *S*_*∞*_ is the point at which all infection ceases, i.e. by setting *I*(*t*_2_) = 0, in Equation (4).

## 6 Results

Since the rules in place at the time of the peak were most similar to level 4, the values of *b* and *a* explored were mostly selected on the basis that they manufacture an erroneously-detected, 99.3 % threshold for level 4. The results, as well as the inputs from which they were obtained, are provided on pages 9 to 11.

## 7 Conclusions

The SARS-CoV-2 pandemic was, very likely, a lot closer to its threshold than the South African data suggested, however, some, possibly external, factor still changed the value of *r*_0_. The author’s opinion is therefore that the contemplated data-deficiencies are unlikely to explain the early peak in the SARS-CoV-2 pandemic on their own. If deficient data did, indeed, play a role, then the more compelling of the two phenomena might be that the true level of infection has been underestimated by a multiplicative factor. It is already a documented phenomenon and it is mathematically less disruptive. The existance of a significant, imperceptible, immune fraction of the population quickly drives *r*_0_ up, admittedly not necessarily to unprecedented levels, however, in so doing, it moves thresholds to even lower values. In contrast, there is very little difference between perceived and true *r*_0_’s, should infections have been underestimated by a factor. It is also already a widely recognised phenomenon that the SARS-CoV-2 virus is often insidious and infection data are incorrect by a substantial factor, whereas a pre-existing, imperceptible immune fraction of the population remains nothing more than a hypothetical construct; a mere thought experiment for the present.

The phenomenon of infections having been underestimated by a multiplicative factor, alone, is unable to comprehensively explain the SARS-CoV-2 peak observed in the South African data, without contemplating improbably-high values. Yet, those improbably-high values (*a* = 60) might be possible in an area like Khayalitsha, considering that a massive 57 % of the inhabitants of Mumbai slum areas tested positive [1]. Revising country-wide infections upward by a single order of magnitude is probably something not too far-fetched and it creates a level-4 threshold of 59.0 % that would be erroneously-detected as being 95.9 %, a perceived value not too far from the actual 99.3 % peak observed (see Table 3). It comes a long way to reconciling the observed with the predicted and it remains an inescapable fact that, in July, *r*_0_ did change fairly abruptly in all the contemplated scenarios of data-deficiency. This is not something suggestive of a threshold.

**Table 3:**
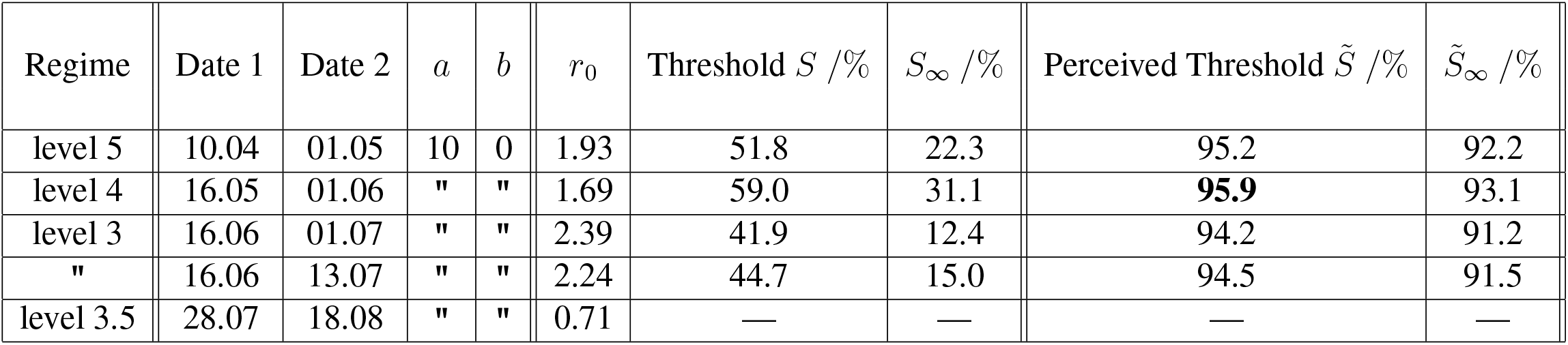
The basic reproduction number, *r*_0_, the consequent threshold, the point at which all infection ceases, *S*_*∞*_, the erroneously detected threshold and the erroneously detected point at which all infection ceases, 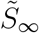.

**Table 4:** The basic reproduction number, *r*_0_, the consequent threshold, the point at which all infection ceases, *S*_*∞*_, the erroneously detected threshold and the erroneously detected point at which all infection ceases,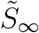.

| Regime | Date 1 | Date 2 | $a$ | $b$ | $r_0$ | Threshold $S$ /% | $S_\infty$ /% | Perceived Threshold $\tilde{S}$ /% | $\tilde{S}_\infty$ /% |
| --- | --- | --- | --- | --- | --- | --- | --- | --- | --- |
| level 5 | 10.04 | 01.05 | 10 | 0.83 | 11.4 | 8.8 | 3.8 | 99.2 | 98.7 |
| level 4 | 16.05 | 01.06 | " | " | 10.2 | 9.8 | 5.0 | **99.3** | 98.8 |
| level 3 | 16.06 | 01.07 | " | " | 15.6 | 6.4 | 1.6 | 98.9 | 98.5 |
| " | 16.06 | 13.07 | " | " | 15.7 | 6.4 | 1.6 | 98.9 | 98.5 |
| level 3.5 | 28.07 | 18.08 | " | " | 8.03 | 12.5 | 5.4 | 99.5 | 98.8 |

**Table 5:** The basic reproduction number, *r*_0_, the consequent threshold, the point at which all infection ceases, *S*_*∞*_, the erroneously detected threshold and the erroneously detected point at which all infection ceases,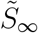.

| Regime | Date 1 | Date 2 | $a$ | $b$ | $r_0$ | Threshold $S$ /% | $S_\infty$ /% | Perceived Threshold $\tilde{S}$ /% | $\tilde{S}_\infty$ /% |
| --- | --- | --- | --- | --- | --- | --- | --- | --- | --- |
| level 5 | 10.04 | 01.05 | 20 | 0.65 | 5.53 | 18.1 | 7.7 | 99.2 | 98.6 |
| level 4 | 16.05 | 01.06 | " | " | 4.94 | 20.3 | 10.3 | **99.3** | 98.8 |
| level 3 | 16.06 | 01.07 | " | " | 7.55 | 13.2 | 3.3 | 98.9 | 98.4 |
| " | 16.06 | 13.07 | " | " | 7.56 | 13.2 | 3.3 | 98.9 | 98.4 |
| level 3.5 | 28.07 | 18.08 | " | " | 3.78 | 26.5 | 11.8 | 99.6 | 98.8 |

**Table 6:** The basic reproduction number, *r*_0_, the consequent threshold, the point at which all infection ceases, *S*_*∞*_, the erroneously detected threshold and the erroneously detected point at which all infection ceases,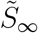.

| Regime | Date 1 | Date 2 | $a$ | $b$ | $r_0$ | Threshold $S$ /% | $S_\infty$ /% | Perceived Threshold $\tilde{S}$ /% | $\tilde{S}_\infty$ /% |
| --- | --- | --- | --- | --- | --- | --- | --- | --- | --- |
| level 5 | 10.04 | 01.05 | 30 | 0.5 | 3.87 | 25.8 | 11.0 | 99.2 | 98.7 |
| level 4 | 16.05 | 01.06 | " | " | 3.46 | 28.9 | 14.7 | **99.3** | 98.8 |
| level 3 | 16.06 | 01.07 | " | " | 5.32 | 18.8 | 4.7 | 99.0 | 98.4 |
| " | 16.06 | 13.07 | " | " | 5.35 | 18.7 | 4.6 | 99.0 | 98.5 |
| level 3.5 | 28.07 | 18.08 | " | " | 2.79 | 35.8 | 15.4 | 99.5 | 98.8 |

**Table 7:** The basic reproduction number, *r*_0_, the consequent threshold, the point at which all infection ceases, *S*_*∞*_, the erroneously detected threshold and the erroneously detected point at which all infection ceases,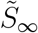.

| Regime | Date 1 | Date 2 | $a$ | $b$ | $r_0$ | Threshold $S$ /% | $S_\infty$ /% | Perceived Threshold $\tilde{S}$ /% | $\tilde{S}_\infty$ /% |
| --- | --- | --- | --- | --- | --- | --- | --- | --- | --- |
| level 5 | 10.04 | 01.05 | 60 | 0 | 1.94 | 51.6 | 22.1 | 99.2 | 98.7 |
| level 4 | 16.05 | 01.06 | " | " | 1.73 | 57.8 | 29.4 | **99.3** | 98.8 |
| level 3 | 16.06 | 01.07 | " | " | 2.66 | 37.6 | 9.3 | 99.0 | 98.5 |
| " | 16.06 | 13.07 | " | " | 2.68 | 37.4 | 9.2 | 99.0 | 98.5 |
| level 3.5 | 28.07 | 18.08 | " | " | 1.40 | 71.6 | 30.8 | 99.5 | 98.8 |

Around the 13th of July, it seems likely that something changed the value of *r*_0_ significantly. Perhaps one criticism of the [12] analysis of lockdown regimes is that it denies the public at large their humanity. The quantification of lockdown regimes means nothing if the rules aren’t complied with. Perhaps, by the 13th of July, the population’s assymptotic compliance had reached the necessary level? Perhaps, too, the intra-household infections that obviously took place early on in the lockdowns, had also run their course? How much genetic drift took place in the preceding months is yet another unanswered question. Perhaps, when the prohibition was reimposed, the seemingly-endless supply of liquor, that had understandably leaked from the stricken hospitality industry, had finally dried up? All these factors could have contributed to lowering the level-3.5 *r*_0_ to below the level-4 value of 1.69, driving the erroneously-detected 95.9 % (Table 3) closer to the 99.3 % peak observed. However, could all of this have driven the 1.69 down as far as 1.08, the *r*_0_-value necessary for the 99.3 % perception? The fact remains that level 3’s *r*_0_ was very much lower than 1.08. It was 0.71 for *a* = 10 and *b* = 0 (Table 3)! This was no threshold.

One external factor possibly worth considering is the change in weather that coincided with the reinstatement of the prohibition and curfew. On the 13th of July, winter arrived in force, imposing a lockdown of its very own. A massive frontal system made its landfall, bringing snow to six provinces of what is, traditionally, a warm country. Braais were called off. Many shack-dwellers were confined to their blankets. Large groups no longer gathered on street corners and outside certain houses to ‘bounce’ liquor and cigrettes. The streets emptied out and finally became deserted. No late-night figure even darted between the throbbing blue houses. People wore their masks to stay warm. Exactly five days later, on the 18th of July, the incidence of SARS-CoV-2 cases peaked (the mean SARS-CoV-2 incubation period is 5.2 days [14]) and the number of active infections followed suit on the 23rd of July. Could something so simple be the explanation? The author is, otherwise, at a loss. It should be borne in mind that the model used is ideally intended for homogenous populations, settlements like Khayalitsha, not really whole countries.

## Data Availability

All data is available online and is listed in the bibliography.

http://www.worldometers.info

http://en.m.wikipedia.org

